# Predicting Robot-Based Tracking Performance After Stroke Using Lesion Features and Clinical Assessments: A Pilot Study

**DOI:** 10.64898/2026.10.28.26366424

**Authors:** Reem M. Azar, Erica L. Waters, Joana Opong-Duah, Rochelle J. Mendonca, Pamela Z. Cacchione, Michelle J. Johnson

## Abstract

**Background:** Following stroke, motor and cognitive impairments often limit functional independence. Personalized rehabilitation requires comprehensive clinical assessments, which are costly and time-consuming. Imaging scans are routinely collected, but their predictive value in determining post-stroke function remains unclear.

**Objectives:** To determine the ability of clinical assessments, lesion features with medical chart information, and a combined clinical and biomedical model to predict kinematic outcomes from a robot-based tracking task in chronic stroke individuals.

**Methods:** Eighteen participants (70±6 years) completed a robot-based tracking task and a battery of motor and cognitive assessments. Three elastic net regression models were developed: 1) clinical assessments only; 2) lesion metrics and chart information only; and 3) a combined model. Models were evaluated using leave-one-subject-out cross-validation, bootstrapping, and permutation testing with false discovery rate correction. Robot-based outcomes included tracking error, normalized distance, velocity error response, and delay.

**Results:** The clinical assessment model significantly predicted tracking error (R²=0.45), delay (R²=0.22), velocity error response (R²=0.18), and normalized distance (R²=0.12). The combined model significantly predicted tracking error (R²=0.37), delay (R²=0.19), and velocity error response (R²=0.11). The imaging model did not significantly predict any outcome. Box and Blocks Test and Color Trails 1 were stable features. The right dorsal attention network was a stable predictor in several combined models.

**Conclusion:** Clinical assessments outperformed lesion features in predicting tracking performance after chronic stroke. Attention- and motor-related measures were the strongest predictors, highlighting the importance of cognitive-motor interactions in function. A larger sample and more granular lesion features may improve prediction from imaging biomarkers.

## 1. Introduction

One in four people over the age of 25 will have a stroke during their life, resulting in approximately 94 million people living with stroke (Feigin et al., 2025). Following stroke, many individuals experience motor and cognitive impairments, which can be detrimental to their quality of life. Specifically, recent studies report post-stroke cognitive impairment at a rate of 60% (El Husseini et al., 2023) and motor impairment at a rate of 61.7% (Clery et al., 2020). High-intensity rehabilitation is essential for increasing functional recovery and independence; however, in low- and middle-income countries (LMIC), access to rehabilitation services is limited (“Global Inequalities in Stroke Rehabilitation: System Capacity, Workforce, and Access in Low- and Middle-Income Countries | Current Physical Medicine and Rehabilitation Reports | Springer Nature Link,” 2026). This need is particularly important as LMICs account for 89% of the global disability adjusted life years due stroke (Feigin et al., 2025). Improving equitable and effective delivery of post-stroke rehabilitation services in low-resource environments requires optimal allocation of rehabilitation services, which may be facilitated by predictive models of rehabilitation needs.

Depending on the size and location of the lesion, a person diagnosed with stroke may develop various motor and cognitive impairments. Imaging information is crucial for predicting recovery as function is influenced by the location of neuroanatomic injury (Jo et al., 2023). Structural brain imaging is routine procedure in post-stroke care, typically performed using Computerized Tomography (CT) or Magnetic Resonance Imaging (MRI). Although MRI-scans offer superior tissue contrast, they are expensive and not widely available in LMICs (Murali et al., 2024). CT-scans, however, are more affordable and more widely available in both low and high-resource settings.

While several studies have investigated the relationship between post-stroke function and neural lesion features using CT-scans, these studies focused on specific regions of interests, rather than exploring the whole brain (Ernst et al., 2018). Focusing on highly specific brain regions does not consider that the brain is deeply interconnected. For example, the premotor cortex (motor domain) relays information between the motor cortex (motor domain) and cognitive processing areas, such as the prefrontal cortex (cognitive domain). Therefore, any disruption in these networks due to stroke may lead to deficits in both motor and cognitive performance. Although few studies have examined how often these deficits co-occur, the available evidence suggests that cognition influences motor performance, and that low or moderate cognitive function can mask otherwise strong motor performance (Bui et al., 2022; Lin et al., 2021). Although information from the acute phase is valuable in clinical decision making, it does not directly inform therapeutic strategies administered in the chronic phase of rehabilitation, when long-term recovery of motor and cognitive abilities occurs. Given that the critical time window for post-stroke recovery extends beyond 12 months, well into the chronic phase of stroke, ongoing assessment throughout post-stroke rehabilitation care is essential (Ballester et al., 2019).

Traditionally, stroke rehabilitation relies on “impairment-level treatments,” which strongly emphasizes motor performance (Shelton FN and Reding MJ, 2001). Administering comprehensive clinical assessment, however, is challenging even in high-resource environments and presents significant challenges in LMICs due to the extensive time required to complete them (∼2-3 hours) and the limited number of experts trained to perform them. While simple self-report measures can be fast and require minimal training to administer, literature supports that stroke patients may not be cognizant of their own disabilities, limiting the efficacy of self-report measures (Hartman-Maeir et al., 2003).

More recently, rehabilitation robots have been proposed as a viable tool for administering clinical assessment, particularly in low-resource environments, with robot systems potentially showing greater sensitivity than standard clinical assessments (Ekechukwu et al., 2020; Logan et al., 2018). Moreover, robot-assisted therapy can deliver real-time feedback, supporting data-driven therapeutic adjustments. Through robot-assisted therapy, personalized neurorehabilitation strategies can be developed from a patient’s current “presenting characteristics” (Jadhav AP et al., 2020). Specifically, robot-assisted therapy can target both motor and cognitive processes, with one study reporting improvements in targeted cognitive domains (Aprile et al., 2020). Despite this preliminary evidence for cognitive recovery, initial robotic intervention strategies are typically based on performance in clinical motor assessments, without considering cognitive domains such as executive function, attention, and working memory. Additionally, because cognitive impairments can mask true motor ability, patients with moderate cognitive impairment and mild motor impairment may be misclassified as having more significant motor impairment (Bui et al., 2022; Lin et al., 2021). This misclassification can distort a patient’s functional profile and lead to suboptimal therapeutic decisions. Thus, relying solely on limited clinical assessment scores assumes a simplistic and unrealistic view of post-stroke recovery, especially if there is not sufficient time or staffing to deliver a comprehensive assessment battery. Furthermore, examining impairment without considering the source, namely brain damage in the form of a lesion, ignores the patient’s true capacity for recovery.

Predicting chronic outcomes is essential, as therapists design interventions based on individual needs, which are often defined by observable motor and cognitive performance. Discovering biomarkers, from non-invasive and accessible technology, that can predict functional recovery in stroke patients is a priority. Literature supports that combining biomarkers (such as imaging) and functional measures to predict recovery is an ideal model [17]. However, existing integrated prognostic models are limited. For example, PREP2 incorporated clinical and imaging measures to predict upper limb recovery, but it relied primarily on measures of overall disability and motor outcome rather than detailed assessments of motor and cognitive performance [17]. To address this gap, we developed models using routinely available information from the medical chart, CT-scan derived lesion metrics, and chronic motor and cognitive assessments to predict kinematic metrics from a complex robot-based task in a retrospective pilot cohort of medically complex patients in the chronic stage of stroke. We compared the performance of these models to one another in order to understand the relative value of different types of clinical information in predicting post-stroke performance.

## 2. Materials and Methods

We use structural imaging, collected from each participant’s electronic health record and processed, along with clinical assessments of motor and cognitive performance to predict robot-based performance of a continuous tracking task. Our primary analysis involves comparing three predictive models: a model of using only clinical assessments (M1), a model using only structural imaging features and information from the medical chart (M2), and a model that combines clinical assessments and structural imaging features (M3). Figure 1 summarizes the methods used in this paper.

**Figure 1:**
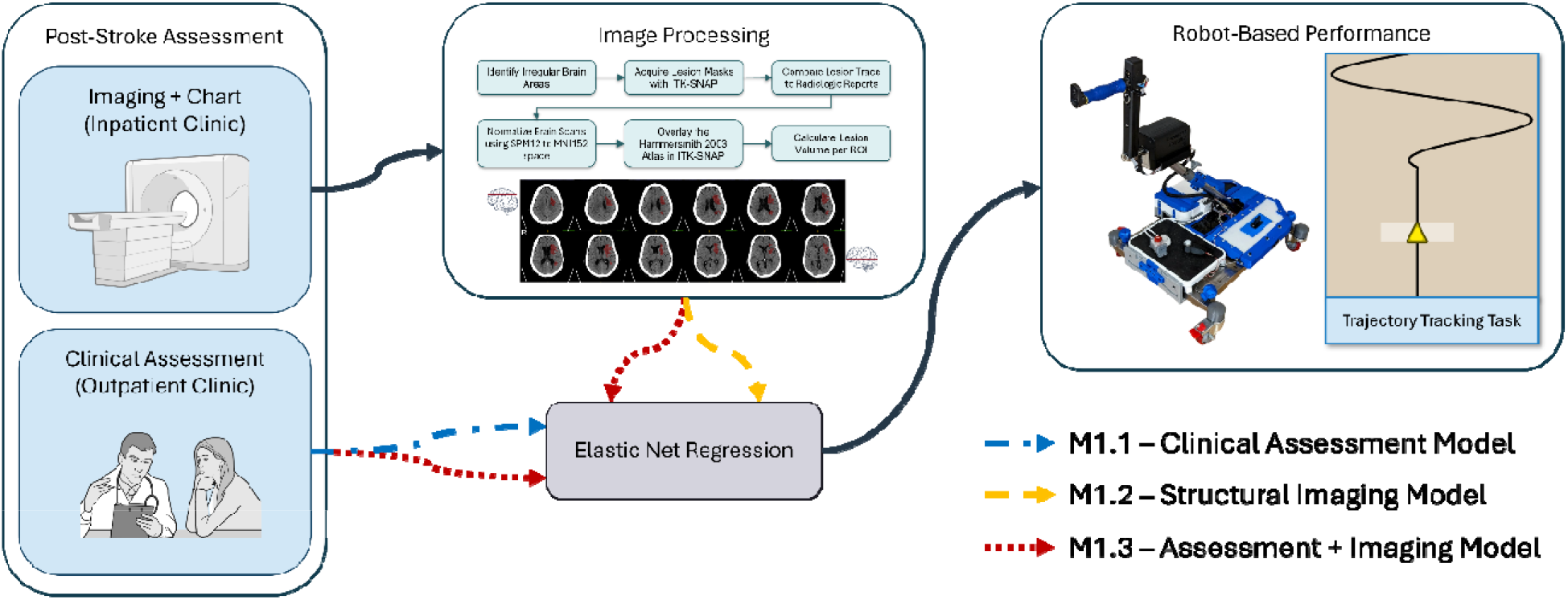
Overview of methods. Workflow linking post-stroke assessment (imaging/chart and clinical) to robot-based tracking performance via elastic net regression. Three models were tested: clinical assessment only (M1), imaging + chart only (M2), and combined (M3).

### 2.1. Participants

The retrospective dataset includes 18 participants (average age 70 ± 6 years) who completed the pre-assessment portion of the Rehabilitation Using Community-Based Affordable Robotic Exercise Systems (Rehab CARES) clinical trial (phase II) (“Rehab CARES In USA,” 2026). This protocol was approved by the Institutional Review Board of the University of Pennsylvania (protocol #853607, approved December 13, 2023). This study is registered at ClinicalTrials.gov under ID: NCT05542121. Table I shows the demographic information for all participants included in this paper.

**Table I:** Participant Demographics and Clinical Characteristics.

| ID | Demographics |  |  |  | Stroke Characteristics |  |  |  |  | Clinical Assessment Scores |  |  |  |  |
| --- | --- | --- | --- | --- | --- | --- | --- | --- | --- | --- | --- | --- | --- | --- |
|  | Age (yrs) | Sex | Ed. (yrs) | Diabetes (Y/N) | Side (L/R/B) | Multiple Strokes (Y/N) | Time Since Stroke (yrs) | HS (Y/N) | Aphasia (Y/N) | BBT-R (blocks) | BBT-L (blocks) | MoCA (score) | MoCA-EF (score) | CT1 (s) |
| 02 | 66-70 | F | 16 | Y | R | Y | 5 | N | Y | 23 | 20 | 13 | 1 | 154 |
| 03 | 61-65 | M | 12 | N | L | Y | 8 | N | Y | 38 | 45 | 16 | 1 | 102 |
| 04 | 71-75 | M | 9-11 | Y | L | N | 34 | Y | N | 39 | 27 | 19 | 2 | 226 |
| 05 | 76-80 | M | 12 | N | B | Y | 15 | Y | Y | 5 | 15 | 9 | 0 | 240 |
| 07 | 71-75 | M | 12 | Y | R | N | 11 | N | Y | 34 | 23 | 18 | 1 | 240 |
| 08 | 71-75 | F | 12 | N | L | N | 1 | Y | Y | 14 | 27 | 10 | 0 | 114 |
| 10 | 66-70 | M | 16 | N | L | N | 9 | Y | Y | 41 | 57 | 14 | 0 | 65 |
| 11 | 66-70 | M | N/A | Y | B | Y | 1 | N | N | 51 | 46 | 25 | 0 | 44 |
| 13 | 66-70 | F | 13-15 | Y | R | Y | 8 | N | N | 52 | 55 | 28 | 4 | 59 |
| 14 | 66-70 | F | 12 | Y | L | N | 3 | N | Y | 47 | 53 | 26 | 5 | 76 |
| 15 | 66-70 | F | 12 | N | B | N | 8 | Y | N | 53 | 49 | 26 | 2 | 135 |
| 17 | 76-80 | F | 13-15 | N | R | N | 2 | Y | N | 48 | 34 | 17 | 3 | 146 |
| 22 | 66-70 | M | 13-15 | N | R | Y | 1 | N | N | 25 | 28 | 22 | 0 | 240 |
| 23 | 61-65 | M | 13-15 | Y | B | Y | 9 | N | N | 31 | 29 | 26 | 5 | 118 |
| 24 | 76-80 | F | 12 | Y | L | Y | 3 | Y | Y | 50 | 46 | 28 | 5 | 130 |
| 26 | 71-75 | M | 12 | Y | B | N | 10 | Y | N | 30 | 29 | 18 | 1 | 190 |
| 28 | 76-80 | F | 12 | Y | B | Y | 1 | Y | Y | 28 | 34 | 24 | 4 | 152 |
| 29 | 56-60 | M | 12 | N | R | N | 1 | N | N | 48 | 1 | 24 | 4 | 37.81 |

**Table II:** Network Description.

| Network | Description |
| --- | --- |
| *Default Mode Network (DMN)* | DMN is involved in internally directed cognitive processes, such as memory retrieval, self-reflection, and future planning. |
| *Dorsal Attention Network (DAN)* | DAN is involved in attention and processing external information during goal-directed behavior. |
| *Salience Network (SAN)* | SAN helps detect important or unexpected stimuli and supports shifting attention toward relevant information. |
| *Visual Network (V1)* | V1 supports visual processing, using visual information to guide behavior. |
| *The Limbic System Network (LIN)* | LIN is involved in emotion and memory, which can indirectly affect both motor and cognitive performance. |
| *Sensorimotor Network (SMN)* | SMN is directly involved in voluntary movement and processing somatosensory feedback. |
| *Central Executive Network (CEN)* | CEN is involved in executive control, including decision-making, working memory, attention, and motor planning. |

Participants were recruited at two sites: Mercy LIFE (Living Independently For Elders) West Philadelphia, a Program of All-inclusive Care for the Elderly (PACE), which provides comprehensive medical and social services to nursing home eligible older adults living in the community, and the Rehabilitation Robotics Lab of University of Pennsylvania, which is located on the first floor of an integrated in-patient and out-patient rehabilitation hospital. Data were collected between February 2024 and August 2025. Inclusion criteria were age 55 or older, an official diagnosis of stroke at least 3 months prior to study start, hemiplegia, and a CT-scan or MR scan completed post-stroke onset. We note that this data set consists of participants who completed the pre-assessment but may have met one or more exclusion criteria that prevented them from completing the full study [18]. All participants gave informed consent.

### 2.2. Lesion Feature Extraction

#### 2.2.1. Regions of Interest

The seven main brain networks will be considered. These networks, defined in Table I, encompass areas across the whole brain and are involved in several distinct functions that require human operation. Many of these brain regions are lateralized, with the right- and left-brain regions covering specific functions. Because sensorimotor control is strongly lateralized, SMN lesion metrics were expressed relative to lesion side as ipsilesional and contralesional measures. In contrast, association networks were retained as left- and right-hemisphere features to preserve potential hemisphere-specific effects related to attention, salience processing, and cognition. Specifically, the right DAN and SAN can affect reaction time and cause neglect, which may lead to global deficits (Menon and Uddin, 2010; Shulman et al., 2010). Damage to the left DAN also causes some issues with neglect and attention that may lead to global deficits, so it was also included (Vossel et al., 2014). Although other networks were lateralized by function, the selected features were more relevant to the task. Hemorrhagic and subcortical strokes were included as a feature due to them being linked to work stroke outcomes (Balk et al., 2026; Liew SL et al., 2022).

#### 2.2.2. Image Processing

Currently, the gold standard for lesion identification is through manual segmentation (Lo et al., 2023) and involves using digital imaging and communications in medicine (DICOM) files. For this paper, a lesion is a hypodense area of irregular tissue on the CT-scan that appears on multiple z-slices (indicating a volumetric loss). Also, in this paper, we used only chronic CT-scans. To manually segment the lesions, we followed the guidelines published by Lo et al (Lo et al., 2023). Segmentations were performed after completing training with the ENIGMA team [24]. Radiological reports served as the ground truth. Expert clinicians were consulted when needed. We used ITK-SNAP, an interactive tool developed for medical imaging segmentation, to create the lesion masks (Horn A et al., 2017; Yushkevich PA and Gerig G, 2017). Our methods for image processing are similar to our previous work (Azar et al., 2025b, 2025a) and detailed further in Appendix A.

### 2.3. Clinical Assessment

Participants completed a battery of cognitive, motor, and quality of life assessments to determine their level of function. The clinical assessments used in this study are based on ones previously correlated with the robotic assessments used [9,29]. To measure motor function, we collected the Box and Blocks Test (BBT): The BBT assesses gross motor performance of the upper limb. Participants are instructed to move as many wooden blocks as possible, one at a time, from one compartment of a box to another within 60 seconds using the tested hand. BBT is commonly used in stroke rehabilitation research, with research suggesting that BBT is influenced by cognitive performance [8,9]. To measure cognition, we collected two cognitive assessments: The Montreal Cognitive Assessment (MoCA): The MoCA is a 10-minute, cognitive screening tool that is used to measure impairment in visuospatial/executive function, naming, memory, attention, language, abstraction, delayed recall, and orientation [30]. Overall, the literature indicates that the MoCA is a valid measure of cognitive impairment in persons with chronic stroke; however, it may have some limitations in measuring the extent of impairment in severely impaired individuals or aphasic individuals. MoCA executive function subscore (MoCA-EF) combines elements from the MoCA assessment that pertain to executive function (backward digit span, trail-making test, word similarities, and F-word listgeneration) and is scored out of five points [31]. Color Trails 1 (CT1) evaluates sustained visual attention and processing speed, requiring participants to connect numbered circles in ascending order while alternating between two colors [32]. Performance is quantified by the time taken to complete each part, with longer completion times indicating poorer performance. Lastly, we collected a self-report measure of well-being through the Neuro-Quality of Life (Neuro-QoL) Satisfaction with Social Roles and Activities.

### 2.4. Robot Assessment

Following the clinical battery, participants completed three robotic assessments: spatial span, n-back, and trajectory tracking. These assessments were initially developed to assess motor and cognitive function in individuals with HIV, stroke, and both HIV and stroke (Bui et al., 2022, 2021). Participants completed the robotic assessments using THEAbot, which is a 1-DoF robot for upper limb rehabilitation developed by Recupero Robotics, LLC. THEAbot is re-configurable with three positions that cover shoulder, elbow, and forearm. For assessment purposes, participants use the robot in position 1 where the robot is positioned to the side of the user with the workspace parallel to the sagittal plane. This movement simulates reach-to-grasp movements and involves flexion/extension of the shoulder and elbow (Meng et al., 2025).

For the purposes of this paper, we focus on the trajectory tracking task, which requires the user to follow the same path for 15 trials lasting 15 seconds each. Performance on continuous tracking tasks, quantified with kinematic outcome measures, have proven effective for identifying post-stroke motor impairment (Feng and Winters, 2009; Jones and Donaldson, 1981). Continuous tracking tasks, however, also involve cognitive domains. Preview tracking tasks in particular have shown correlations with cognitive assessments related to executive function and attention in addition to motor assessments related to motor impairment and motor activity (Bui et al., 2022, 2021). Continuous preview tracking tasks allow for constant action monitoring and better imitate human behavior compared to discrete tasks (Pereira et al., 2017). Such tasks are highly relevant to activities of daily living, e.g. driving, which require motor and cognitive domains (Jones and Donaldson, 1981). Trajectory tracking, therefore, is ideal to study multi-network interaction in the brain because it involves a complex task with both cognitive and motor inputs, and it is highly relevant to activities of daily living.

### 2.5. Predictive Modeling Procedure

To predict kinematic outcomes and simulation model parameters, we utilized elastic net regression, a form of penalized regression that combines L1 and L2 penalties, encouraging sparsity by setting some coefficients to zero while also accounting for co-linearity in predictors through a bias-variance trade-off (“Regularization and Variable Selection via the Elastic Net on JSTOR,” n.d.). Two parameters, λ and α, define the model, where λ is the penalty strength parameter and α is the ratio between L1 and L2 penalties (α = 0 for ridge regression; α = 1 for lasso regression).

#### 2.5.1. Outcome Variables

A total of six outcome variables were identified: three kinematic metrics and three simulation model parameter metrics. The kinematic metrics included root mean square tracking error (RMSE), normalized distance (ND), and spectral arc length (SPARC). RMSE represents the square root of the mean squared tracking error where a higher value corresponds to worse performance (Bui et al., 2021). ND represents the total distance traveled divided by the distance required to exactly follow the target; a value of one represents moving as much as the task requires while values above and below one represents moving more and less, respectively (Bui et al., 2021). SPARC is a measure of smoothness defined as the arc length of the Fourier magnitude spectrum of the speed profile; a more negative value corresponds to less smooth movement (Balasubramanian et al., 2015).

For simulation model parameters, we fit a feedback control simulation model, which is defined in Appendix B and in our previous work (Waters et al., 2025; Waters and Johnson, 2025). From this simulation model, we identified parameters including Kv, Delay, and Bias. Kv represents the gain on error in velocity; a greater value corresponds to a larger response to velocity error. Delay represents the average time shift that maximizes cross-correlation between the participant’s trajectory and the target’s trajectory. A higher value corresponds to greater delay in response. Bias represents the average median value of human position where the median position for the target is zero. The absolute value of Bias was used such that a bias to either side is treated the same.

#### 2.5.2. Predictor Features

Three different feature matrices were constructed for the three different model types. For the M1 Clinical Assessment Model, we identified four candidate predictors: CT1, MoCA-EF, BBT, and Neuro-QoL Satisfaction. For the M2 Imaging + Chart Model, we identified four features from the chart and ten features derived from the imaging analysis. For chart features, diabetes, aphasia, and hemorrhagic stroke were collected and treated as binary variables while age was a continuous variable. These features were selected because they had a good distribution in the available population, were available in the medical chart, and had evidence of a relationship to the selected task (Balk et al., 2026) (Xu et al., 2021) (Lau et al., 2019). For image derived features, we selected total lesion volume in the subcortical region as well as lesion volume in specific brain networks. For the M3 Combined Model, we used all 18 features from both previous models.

#### 2.5.3. Model Tuning and Assessment

The three model types (M1 Clinical Assessment Model; M2 Imaging + Chart Model; M3 Combined Model) were fit to each of the six outcome metrics for a total of 18 models. To improve interpretability, all outcome variables were sign-oriented such that a more positive value indicated worse performance; all candidate predictor features were oriented such that a more positive value corresponded to expected worse outcome. When fitting the model, we required all coefficients to be positive, ensuring that all predictor-outcome relationships were biologically justifiable, e.g., greater lesion volume leading to more deficit. Prior to fitting each model, we z-score normalized the continuous candidate predictors in the feature matrix.

Model performance was leave-one-subject-out (LOSO) cross-validated and assessed with the coefficient of determination (R^2^). Within each cross-validation fold, hyperparameters were tuned using LOSO cross-validation to select the best λ and α values. We restricted α values to favor Lasso regression (0.5 to 0.9 with step size 0.1) in order to encourage feature sparsity. The set of parameters (λ, α) with the lowest cross-validation error were selected as the best performing model within each fold. Each regression model was then bootstrapped (n = 10,000) to generate the 95% confidence interval around the observed R^2^. Finally, permutation testing (n = 10,000) was performed on each model to test the likelihood that a given cross-validated R^2^ could be achieved at random with a null model. Significance p-values from permutation tests were adjusted for multiple comparisons across models using the Benjamini– Hochberg false discovery rate (FDR) procedure; all reported p-values are FDR-adjusted. All analyses were carried out using Python scikit-learn library.

## 3. Results

### 3.1. Lesion Metrics

Lesions volumes were normalized within networks for proper comparison across groups. Two participants did not have available CT scans, so their MR scans were used. All participants had lesion involvement in the CEN and DMN, 94.4% patients had involvement of the DAN, 88.9% had involvement in the SAN and LIN, 83.3% had involvement in the SMN, and 50% had involvement in V1 (Figure C.2). Eight participants had involvement across all seven networks, six had involvement in six networks, two had involvement in five networks, one had involvement in four networks, and one had involvement in three networks. Although network involvement was represented, the lesion volume for these areas was variable.

### 3.2. Model Performance

Figure 2 summarizes the performance of all models across the six outcomes. After adjusting for multiple hypothesis testing, eight models achieved significance. Specifically, the assessment models were significant for RMSE, ND, Kv, and Delay. The combined model was likewise significant for RMSE, Kv, and Delay but not ND. Table II displays performance results for a l models.

**Figure 2:**
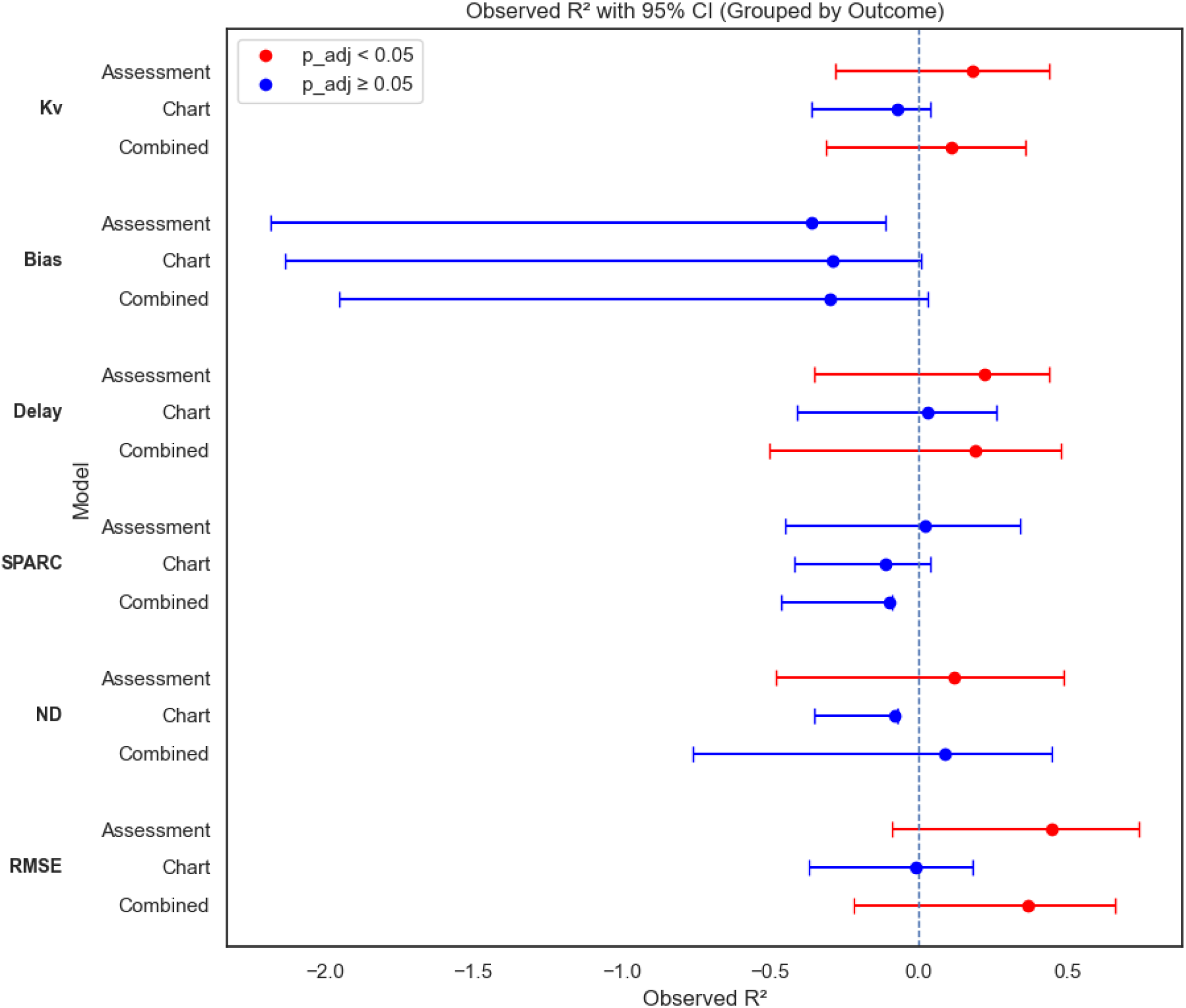
Observed Correlation Coefficient by Outcome for the 3 Models. Observed R^2^ values with 95% confidence intervals for elastic net regression models of six outcome metrics. A red marker indicates that the model performed significantly better than random chance (FDR adjusted p < 0.05).

**Table II:** Model Performance.

| Model | Metric* | $\lambda$ [IQR] | $\alpha$ [IQR] | $R^2$ [95% CI] | p (adj.) | Sig. |
| --- | --- | --- | --- | --- | --- | --- |
| M3 –<br>Combined | RMSE | 0.81 [0.68, 0.90] | 0.6 [0.6, 0.7] | 0.37 [-0.22, 0.66] | 0.003 | ** |
|  | ND | 0.06 [0.05, 0.06] | 0.5 [0.5, 0.5] | 0.09 [-0.76, 0.45] | 0.068 |  |
|  | SPARC | 1.16 [1.06, 2.32] | 0.5 [0.5, 0.5] | -0.10 [-0.46, 0.09] | 0.291 |  |
|  | Delay | 0.07 [0.06, 0.08] | 0.5 [0.5, 0.5] | 0.19 [-0.50, 0.48] | 0.030 | * |
|  | Bias | 1.04 [0.90, 1.21] | 0.5 [0.5, 0.5] | -0.30 [-1.95, 0.03] | 0.109 |  |
|  | Kv | 1.2 [1.15, 1.27] | 0.5 [0.5, 0.75] | 0.11 [-0.31, 0.36] | 0.013 | * |
| M2 – Chart<br>+ Imaging | RMSE | 1.11 [1.05, 1.16] | 0.9 [0.83, 0.9] | -0.01 [-0.37, 0.18] | 0.089 |  |
|  | ND | 0.17 [0.15, 0.18] | 0.5 [0.5, 0.5] | -0.08 [-0.35, -0.07] | 0.768 |  |
|  | SPARC | 2.10 [1.98, 2.14] | 0.5 [0.5, 0.5] | -0.11 [-0.42, -0.04] | 0.799 |  |
|  | Delay | 0.06 [0.05, 0.08] | 0.9 [0.53, 0.9] | 0.03 [-0.41, 0.26] | 0.073 |  |
|  | Bias | 0.73 [0.72, 0.81] | 0.5 [0.5, 0.5] | -0.29 [-2.13, 0.01] | 0.106 |  |
|  | Kv | 0.86 [0.76, 0.88] | 0.9 [0.8, 0.9] | -0.07 [-0.36, 0.04] | 0.092 |  |
| M1 –<br>Clinical<br>Assessment | RMSE | 0.27 [0.25, 0.31] | 0.5 [0.5, 0.5] | 0.45 [-0.09, 0.74] | 0.003 | ** |
|  | ND | 0.04 [0.03, 0.04] | 0.5 [0.5, 0.5] | 0.12 [-0.48, 0.49] | 0.033 | * |
|  | SPARC | 0.32 [0.28, 0.34] | 0.5 [0.5, 0.5] | 0.02 [-0.45, 0.34] | 0.073 |  |
|  | Delay | 0.02 [0.02, 0.03] | 0.5 [0.5, 0.5] | 0.22 [-0.35, 0.44] | 0.010 | * |
|  | Bias | 4.08 [3.88, 4.30] | 0.5 [0.5, 0.5] | -0.36 [-2.18, -0.11] | 0.786 |  |
|  | Kv | 0.60 [0.58, 0.67] | 0.5 [0.5, 0.5] | 0.18 [-0.28, 0.44] | 0.010 | * |

\*Metrics: RMSE (Root Mean Square Error), measure of tracking accuracy; ND (normalized distance), measure of movement efficiency; SPARC (spectral arc length), measure of movement smoothness; Delay, measure of lag (in seconds) behind the target during tracking; Bias, measure of spatial shift in range-of-motion during tracking relative to the median task position; Kv, measure of response to error in velocity during tracking

RMSE was the best predicted outcome with moderate predictive performance and confidence intervals skewed highly positive for both the assessment and combined models with both models achieving similar performance. The chart model for RMSE, however, performed much worse with a negative R^2^ and negative skewed confidence interval.

Delay and Kv were the next best predicted outcomes with mild predictive performance and confidence intervals roughly centered on zero. Assessment and combined models achieve similar performance for both Delay and Kv. The chart model performed poorly for Kv with only slightly better performance for Delay. ND achieved significance for the assessment but not the combined model or chart model.

Neither SPARC nor Bias achieved significance in any of the models. The SPARC assessment model performed the best of the three, while the chart and combined models demonstrated poor performance with confidence intervals skewed negative. Bias performed the worst overall with all models demonstrating poor performance and confidence intervals with a highly negative skew.

### 3.3. Selected Features

Figure 3 shows the features selected as predictors for each model. Features were highly stable across cross-validation folds with selected features achieving a median selection frequency of 64%, IQR [6%, 100%] across all models and a median selection frequency of 97%, IQR [6%, 100%] among the models that performed significantly better than the null (Table D.1). High α values (≥0.5) biased the models towards Lasso regression and helped to keep total number of selected features to a minimum with models selecting between 0 and 4 stable features. Stable features were defined as features with a selection frequency of 50% or higher.

**Figure 3:**
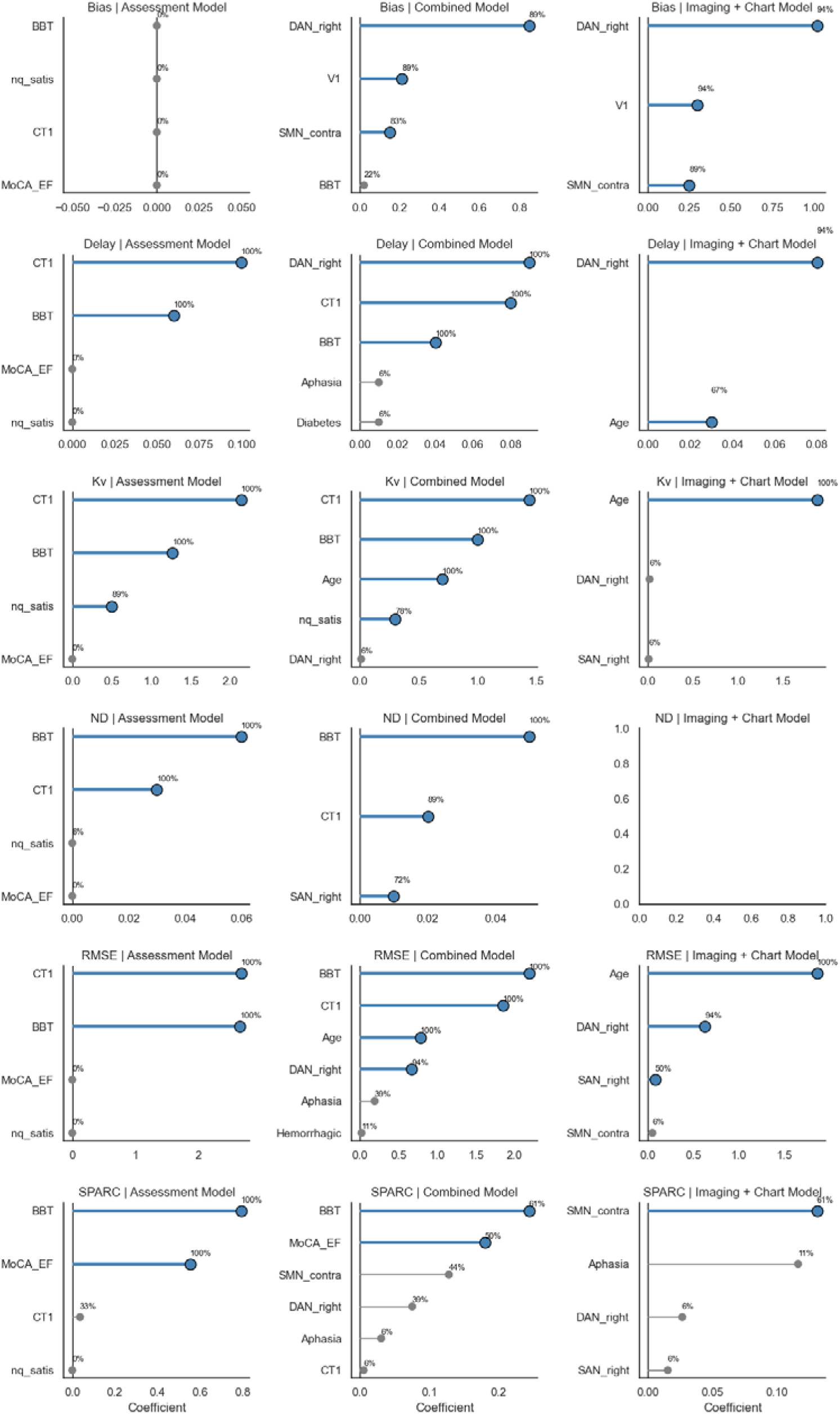
Feature Selection. Features selected by each model with selection frequency and mean coefficient value. Mean coefficient value is represented on the x-axis. Selection frequency is denoted to the right of each value.

Looking at stable features for the significant models, the assessment models for RMSE, Delay, Kv, and ND consistently selected CT1 and BBT (100% selection frequency) with the Kv model also selecting Neuro-QoL satisfaction (89% selection frequency). The combined models for RMSE, Delay, and Kv preserved these stable features, maintaining universal selection of CT1 and BBT across all three significant models (100% selection frequency) and high selection of Neuro-QoL satisfaction in the Kv model (78% selection frequency). Likewise, age was consistently selected for the RMSE and Kv combined models (100% selection frequency) while DAN right was selected for RMSE and Delay (94% and 100% selection frequencies, respectively). Although the chart models for RMSE, Delay, and Kv were not significant, we note they selected similar stable features as the combined models, with age selected across all models (100% selection frequency for RMSE and Kv, 94% selection frequency for Delay) and DAN right consistently selected for RMSE and Delay (94% selection frequency for both).

## 4. Discussion

In this paper, we used medical chart information, lesion metrics, and motor and cognitive assessment metrics to predict kinematic metrics from a motor tracking task. While some studies have assessed the predictive value of lesion metrics on behavioral outcomes, these studies primarily predict a dichotomized outcome (good vs. severe) or overall disability status. To our knowledge, this study is the first study to evaluate a biomarker model, clinical model, and combined model as predictors of continuous kinematic markers from a robotic assessment.

### 4.1. Clinical Assessments Outperform Neural Lesion Features

Research-standard motor and cognitive assessments outperformed the imaging model and performed similarly to the combined model. Specifically, the clinical assessment model had significant results in predicting Kv, Delay, ND, and RMSE, whereas the combined model only had significant results in Kv, Delay, and RMSE. These results contrast with prior reports that combined biomarker model outperforms individual models; however, prior studies rely heavily on overall measurements of disability (similarly to assessment performed in treatment settings), classification models, and binary outcomes (Brzus et al., 2025; Stinear et al., 2017). Furthermore, the similar performance of the combined and clinical model suggests that the clinical features may capture most of the variance relevant to the robotic task and are the gold standard for understanding motor and cognitive behavior post-stroke.

Unlike the clinical assessment model, the chart model, which included neuroimaging features and co-morbidities, did not significantly predict any outcome variable. One explanation for the poor performance in the chart/imaging model is the small sample size. Stroke-related lesions are highly heterogeneous. In the small, diverse cohort presented here, we noted that many lesion networks were unevenly distributed (Figure C.2) leading to network feature representation in only a subset of the cohort. Given the small sample size, we limited the lesion mapping to the seven main brain networks. Imaging features at this level of granularity, however, represent general function and may may not directly measure the specific processes involved in the robotic task. For example, the trajectory tracking task required visual processing, sustained attention, motor planning, error correction, and motor control. Although certain regions within the networks deal with these functions, the entire network encompasses task irrelevant regions. In contrast, CT1 and BBT directly assess relevant processes like sustained attention and motor speed. Finally, structural lesion mapping does not consider neuroplasticity. Non-contrast structural imaging cannot capture plastic changes in the brain, as neuroplasticity usually involves synaptic reorganization (Patel et al., 2025). Overtime, the relationship between lesion location and behavior may become less direct as patients recover and plasticity occurs. Our cohort was well into the chronic phase of stroke (7 years ± 8 years post stroke) and therefore had already experienced plasticity. In the acute and sub-acute phases, however, it may be possible to more directly predict performance.

Overall, these findings suggest that targeted clinical assessment metrics were sufficient to predict some kinematic outcomes on a preview tracking task and that medical chart data did not add predictive value. It is possible that neuroimaging information would be useful in post-stroke prediction for larger sample sizes and with more granular features, but we note that the structural imaging approach described here will not account for neuroplasticity. Nevertheless, when combined with up-to-date clinical assessments, structural lesion mapping could help to better understand chronic post-stroke impairment.

### 4.2. Cognition, Particularly Attention, is Predictive of Preview Tracking Performance

Motor performance is task relevant, as demonstrated through the consistent selection of BBT across significant models. The consistent selection of CT1 across significant models (Kv, RMSE, ND, and Delay), however, validates that the robotic task is not purely motor, but rather, depends on sustained visual attention and processing speed. These results support previous work from our lab that showed that MoCA-EF was related to RMSE and ND in the trajectory tracking task, while CT1 was related to ND (Bui et al., 2022, 2021). MoCA is highly variable to aphasia and MoCA-EF has low granularity being scored out of five points, which may explain why it was not significant in this study (Benaim et al., 2015; Chiti and Pantoni, 2014). We also note that the task explored here is almost twice as fast as the task presented in our previous work. The faster speed may explain why attention rather than executive emerged as a significant predictor.

Interestingly, the RMSE models showed similar sized coefficients for CT1 and BBT while ND models showed larger coefficients for BBT relative to CT1. The Delay and Kv models, however, showed greater coefficients for CT1 relative to BBT. Furthermore, the Kv model showed Neuro-QoL Satisfaction as a small but stable predictor. Together these results suggest that the model-derived parameters may be more sensitive to cognitive deficits than traditional kinematic metrics. While RMSE and ND represent overall task performance, past research has shown that these metrics do not distinguish chronic stroke participants with moderate motor impairment and low cognitive impairment from those with low motor impairment and moderate cognitive impairment (Bui et al., 2022). In other words, cognitive impairment can mask motor ability when assessed with standard kinematic metrics. The model parameters, Delay and Kv, however, help define motor control and serve as proxies for specific elements of tracking behavior, i.e. timing of response and response to error. These metrics, therefore, may help to better characterize tracking performance, reporting the source of error, e.g. delay or insufficient response.

Although the chart model was unable to predict robotic outcomes independently, stable selected features support the importance of attention in the selected task. Specifically, right DAN emerged as a stable feature in the significant combined models, with a selection frequency of 94% for RMSE and 100% for Delay. Outcomes such as Delay and RMSE reflect movement accuracy and timing, and literature supports the right DAN’s role in relevant domains like spatial attention and visuospatial processing (Menon and Uddin, 2010; Shulman et al., 2010). These findings support that a combined model may allow for emergence of neuroimaging features, although these features alone may not be enough to produce a significant prediction model at this sample size and feature dimensionality.

### 4.3. Clinical Implications

In this study, we demonstrated that research-standard assessments outperform neuroimaging and medical chart data in predicting kinematic outcomes in a chronic stroke population. However, chronic assessments of behavior are not acquired frequently as many stroke patients are discharged at that point (Shahid et al., 2023). Even in the acute phase, we found that many patients in our study did not have motor or cognitive assessments: two patients had a MoCA, seven had an NIHSS (National Institute of Health Stroke Severity Scale), and none had a Barthel Index (measurement of functional independence). Thus, building a predictive model of chronic behavior using research-standard assessments may not be realistic.

CT imaging is routinely collected post-stroke, even in low-resource environments. Information from these scans may be used to understand recovery and design better acute and chronic therapeutic interventions. Specifically, motor and cognitive interventions early on may lead to better rehabilitation outcomes. Despite neuroplastic reorganization, lesion location can limit recovery. For example, brainstem strokes, although small, are the most devastating type of stroke and hardest to recover. Thus, knowing lesion location early on may be valuable to understanding recovery potential.

Traditionally, robot-assisted therapies and conventional therapies have targeted motor impairment, but cognitive impairment is closely linked to motor performance post-stroke (Bui et al., 2022; Lin et al., 2021). This complex interaction between motor and cognitive impairments necessitates comprehensive motor and cognitive assessment to identify optimal candidate and treatment strategies. Quantitative robotic assessments can improve post-stroke assessment by providing combined motor and cognitive assessments, automatic data collection, and quick, objective functional measures.

## 5. Conclusion

This preliminary pilot study highlights the value of motor and cognitive assessments combined with neuroimaging. We demonstrate that cognitive function, particularly attention, along with motor activity are important predictors of post-stroke tracking performance. With a larger dataset, it may be possible to include more features, such as more task-specific networks or smaller, targeted regions of interest. The findings in this paper, although conservative, provide a promising foundation for using objective, affordable robotic technology to better characterize post-stroke impairment.

## Credit Author Statement

Reem Azar: Conceptualization, Methodology, Formal Analysis, Investigation, Data Curation, Writing - Original Draft, Project Administration, Funding Acquisition. Erica Waters: Conceptualization, Methodology, Software, Formal Analysis, Data Curation, Writing - Original Draft, Visualization, Funding Acquisition. Joana Opon-Duah: Investigation. Rochelle Mendonca: Supervision, Writing - Review & Editing. Pamela Cacchione: Supervision, Project Administration, Writing - Review & Editing. Michelle Johnson: Conceptualization, Resources, Writing - Review & Editing, Supervision, Project Administration, Funding Acquisition.

## Data Availability

Per the guidelines of this journal, sharing the data that support the findings in this paper is required unless it is not possible. The data cannot be made publicly available because they contain sensitive medical and clinical information from human subjects, including information from neuroimaging scans. Public sharing of the data will be kept to a minimum. De-identified data may be made available upon reasonable request and approval from the institutional review board that at the University of Pennsylvania.

## Acknowledgement

We want to thank Brian Smart, OTR/L, Kelly Conway, RN, OTR/L, Carol Wamsley, DPT, and Greg Holt, OTR/L for conducting patient assessments. We also thank Alwyn Johnson, Samuel Gaardsmoe, and Saibaba Nargund at Recupero Robotics, LLC for designing and supporting the robot systems and enAblegames™ for implementing the robotic assessments and software architecture. Finally, we thank Mercy LIFE West Philadelphia for providing clinical space and assistance with recruitment and the participants for their enthusiasm.

## Declaration of generative AI and AI-assisted technologies in the manuscript preparation process

During the preparation of this work, the authors used ChatGPT and Microsoft Copilot in order to brainstorm, literature search, write analysis code, and review grammar for clarity. After using this tool/service, the authors reviewed and edited the content as needed and take full responsibility for the content of the published article.

## References

Aprile, I., Germanotta, M., Cruciani, A., Loreti, S., Pecchioli, C., Cecchi, F., Montesano, A., Galeri, S., Diverio, M., Falsini, C., Speranza, G., Langone, E., Papadopoulou, D., Padua, L., Carrozza, M.C., FDG Robotic Rehabilitation Group, 2020. Upper Limb Robotic Rehabilitation After Stroke: A Multicenter, Randomized Clinical Trial. J. Neurol. Phys. Ther. JNPT 44, 3–14. 10.1097/NPT.0000000000000295

Azar, R.M., Sobotka, N.S., Cacchione, P., Mendonca, R.J., Johnson, M.J., 2025a. Evaluating the Relationship Between Motor and Cognitive Impairment and Lesion Size and Location Post-Stroke Using CT Scans*, in: 2025 International Conference On Rehabilitation Robotics (ICORR). Presented at the 2025 International Conference On Rehabilitation Robotics (ICORR), pp. 1467–1472. 10.1109/ICORR66766.2025.11063052

Azar, R.M., Waters, E.L., Opong-Duah, J., Cacchione, P.Z., Johnson, M.J., 2025b. Modeling Post-Stroke Motor Behavior from Structural Lesion Networks: A Preliminary Study with Robot-Based Assessments. Proceedings of the IEEE International Conference on Neural Engineering (NER).

Balasubramanian, S., Melendez-Calderon, A., Roby-Brami, A., Burdet, E., 2015. On the analysis of movement smoothness. J. NeuroEngineering Rehabil. 12, 112. 10.1186/s12984-015-0090-9

Balk, S., Siller, T., Sprügel, M.I., Haupenthal, D., Kölbl, K., Hock, S., Heinze, D., Engelhorn, T., Kallmünzer, B., Schwab, S., Huttner, H.B., Kuramatsu, J.B., Sembill, J.A., 2026. Functional outcome, five-year survival and burden of disease after size- and location-matched hemorrhagic versus ischemic stroke. Neurol. Res. Pract. 8, 6. 10.1186/s42466-026-00456-w

Ballester, B.R., Maier, M., Duff, A., Cameirão, M., Bermúdez, S., Duarte, E., Cuxart, A., Rodríguez, S., San Segundo Mozo, R.M., Verschure, P.F.M.J., 2019. A critical time window for recovery extends beyond one-year post-stroke. J. Neurophysiol. 122, 350–357. 10.1152/jn.00762.2018

Benaim, C., Barnay, J.L., Wauquiez, G., Bonnin-Koang, H.Y., Anquetil, C., Pérennou, D., Piscicelli, C., Lucas-Pineau, B., Muja, L., le Stunff, E., de Boissezon, X., Terracol, C., Rousseaux, M., Bejot, Y., Antoine, D., Binquet, C., Devilliers, H., 2015. The Cognitive Assessment scale for Stroke Patients (CASP) vs. MMSE and MoCA in non-aphasic hemispheric stroke patients. Ann. Phys. Rehabil. Med. 58, 78–85. 10.1016/j.rehab.2014.12.001

Brzus, M., Griffis, J., Riley, C.J., Bruss, J., Shea, C., Johnson, H.J., Boes, A.D., 2025. A Clinical Neuroimaging Platform for Rapid, Automated Lesion Detection and Personalized Post-Stroke Outcome Prediction. medRxiv 2025.05.09.25327310. 10.1101/2025.05.09.25327310

Bui, K.D., Lyn, B., Roland, M., Wamsley, C.A., Mendonca, R., Johnson, M.J., 2022. The Impact of Cognitive Impairment on Robot-Based Upper-Limb Motor Assessment in Chronic Stroke. Neurorehabil. Neural Repair 36, 587–595. 10.1177/15459683221110892

Bui, K.D., Wamsley, C.A., Johnson, M.J., 2021. Robot-based evaluation of motor and cognitive impairment in people living with HIV and HIV-associated stroke. Neurorehabil Neural Repair.

Chiti, G., Pantoni, L., 2014. Use of Montreal Cognitive Assessment in patients with stroke. Stroke 45, 3135–3140. 10.1161/STROKEAHA.114.004590

Clery, A., Bhalla, A., Rudd, A.G., Wolfe, C.D.A., Wang, Y., 2020. Trends in prevalence of acute stroke impairments: A population-based cohort study using the South London Stroke Register. PLOS Med. 17, e1003366. 10.1371/journal.pmed.1003366

Ekechukwu, E.N.D., Olowoyo, P., Nwankwo, K.O., Olaleye, O.A., Ogbodo, V.E., Hamzat, T.K., Owolabi, M.O., 2020. Pragmatic Solutions for Stroke Recovery and Improved Quality of Life in Low- and Middle-Income Countries-A Systematic Review. Front Neurol. 10.3389/fneur.2020.00337

El Husseini, N., Katzan, I.L., Rost, N.S., Blake, M.L., Byun, E., Pendlebury, S.T., Aparicio, H.J., Marquine, M.J., Gottesman, R.F., Smith, E.E., on behalf of the American Heart Association Stroke Council; Council on Cardiovascular and Stroke Nursing; Council on Cardiovascular Radiology and Intervention; Council on Hypertension; and Council on Lifestyle and Cardiometabolic Health, 2023. Cognitive Impairment After Ischemic and Hemorrhagic Stroke: A Scientific Statement From the American Heart Association/American Stroke Association. Stroke 54, e272–e291. 10.1161/STR.0000000000000430

Ernst, M., Boers, A.M.M., Forkert, N.D., Berkhemer, O.A., Roos, Y.B., Dippel, D.W.J., van der Lugt, A., van Oostenbrugge, R.J., van Zwam, W.H., Vettorazzi, E., Fiehler, J., Marquering, H.A., Majoie, C.B.L.M., Gellissen, S., MR CLEAN trial investigators (www.mrclean-trial.org), 2018. Impact of Ischemic Lesion Location on the mRS Score in Patients with Ischemic Stroke: A Voxel-Based Approach. AJNR Am. J. Neuroradiol. 39, 1989–1994. 10.3174/ajnr.A5821

Feigin, V.L., Brainin, M., Norrving, B., Martins, S.O., Pandian, J., Lindsay, P., F Grupper, M., Rautalin, I., 2025. World Stroke Organization: Global Stroke Fact Sheet 2025. Int. J. Stroke 20, 132–144. 10.1177/17474930241308142

Feng, X., Winters, J.M., 2009. A pilot study evaluating use of a computer-assisted neurorehabilitation platform for upper-extremity stroke assessment. J. Neuroengineering Rehabil. 6, 15. 10.1186/1743-0003-6-15

Global Inequalities in Stroke Rehabilitation: System Capacity, Workforce, and Access in Low- and Middle-Income Countries | Current Physical Medicine and Rehabilitation Reports | Springer Nature Link [WWW Document], 2026. URL https://link.springer.com/article/10.1007/s40141-026-00535-8 (accessed 7.1.26).

Hartman-Maeir, A., Soroker, N., Oman, S.D., Katz, N., 2003. Awareness of disabilities in stroke rehabilitation--a clinical trial. Disabil. Rehabil. 25, 35–44.

Horn A, Kuhn AA, Merkl A, Shih L, Alterman R, Fox M, 2017. Probabilistic conversion of neurosurgical DBS electrode coordinates into MNI space. Neuroimage 150, 395–404. 10.1016/j.neuroimage.2017.02.004

Jadhav AP, Desai SM, Liebeskind DS, Wechsler LR, 2020. Neuroimaging of Acute Stroke. Neurol Clin 38, 185–199. 10.1016/j.ncl.2019.09.004

Jo, H., Kim, C., Gwon, D., Lee, Jaeho, Lee, Joonwon, Park, K.M., Park, S., 2023. Combining clinical and imaging data for predicting functional outcomes after acute ischemic stroke: an automated machine learning approach. Sci. Rep. 13, 16926. 10.1038/s41598-023-44201-8

Jones, R.D., Donaldson, I.M., 1981. Measurement of integrated sensory-motor function following brain damage by a computerized preview tracking task. Int. Rehabil. Med. 3, 71–83. 10.3109/03790798109166740

Lau, L., Lew, J., Borschmann, K., Thijs, V., Ekinci, E.I., 2019. Prevalence of diabetes and its effects on stroke outcomes: A meta-analysis and literature review. J. Diabetes Investig. 10, 780–792. 10.1111/jdi.12932

Liew SL, Lo BP, Donnelly MR, Zavaliangos-Petropulu A, Jeong JN, Barisano G, Hutton A, Simon JP, Juliano JM, Suri A, Wang Z, Abdullah A, Kim J, Ard T, Banaj N, Borich MR, Boyd LA, Brodtmann A, Buetefisch CM, Cao L, Cassidy JM, Ciullo V, Conforto AB, Cramer SC, Dacosta-Aguayo R, de la Rosa E, Domin M, Dula AN, Feng W, Franco AR, Geranmayeh F, Gramfort A, Gregory CM, Hanlon CA, Hordacre BG, Kautz SA, Khlif MS, Kim H, Kirschke JS, Liu J, Lotze M, MacIntosh BJ, Mataro M, Mohamed FB, Nordvik JE, Park G, Pienta A, Piras F, Redman SM, Revill KP, Reyes M, Robertson AD, Seo NJ, Soekadar SR, Spalletta G, Sweet A, Telenczuk M, Thielman G, Westlye LT, Winstein CJ, Wittenberg GF, Wong KA, Yu C, 2022. A large, curated, open-source stroke neuroimaging dataset to improve lesion segmentation algorithms. Sci Data 9, 320. 10.1038/s41597-022-01401-7

Lin, D.J., Erler, K.S., Snider, S.B., Bonkhoff, A.K., DiCarlo, J.A., Lam, N., Ranford, J., Parlman, K., Cohen, A., Freeburn, J., Finklestein, S.P., Schwamm, L.H., Hochberg, L.R., Cramer, S.C., 2021. Cognitive Demands Influence Upper Extremity Motor Performance During Recovery From Acute Stroke. Neurology. 10.1212/WNL.0000000000011992

Lo, B.P., Donnelly, M.R., Barisano, G., Liew, S.-L., 2023. A standardized protocol for manually segmenting stroke lesions on high-resolution T1-weighted MR images. Front. Neuroimaging 1. 10.3389/fnimg.2022.1098604

Logan, L.M., Semrau, J.A., Debert, C.T., Kenzie, J.M., Scott, S.H., Dukelow, S.P., 2018. Using Robotics to Quantify Impairments in Sensorimotor Ability, Visuospatial Attention, Working Memory, and Executive Function After Traumatic Brain Injury. J. Head Trauma Rehabil. 33, E61. 10.1097/HTR.0000000000000349

Meng, N.Y., Waters, E.L., Lin, G., Gaardsmoe, S., Johnson, A., Hallock, L.A., Mendonca, R.J., Cacchione, P.Z., Johnson, M.J., 2025. Analysis of Induced Muscle Activations by an Affordable, Reconfigurable Robot for Comprehensive Post-Stroke Rehabilitation, in: 2025 International Conference On Rehabilitation Robotics (ICORR). Presented at the 2025 International Conference On Rehabilitation Robotics (ICORR), pp. 1361–1367. 10.1109/ICORR66766.2025.11063060

Menon, V., Uddin, L.Q., 2010. Saliency, switching, attention and control: a network model of insula function. Brain Struct. Funct. 214, 655–667. 10.1007/s00429-010-0262-0

Murali, S., Ding, H., Adedeji, F., Qin, C., Obungoloch, J., Asllani, I., Anazodo, U., Ntusi, N.A.B., Mammen, R., Niendorf, T., Adeleke, S., 2024. Bringing MRI to low- and middle-income countries: Directions, challenges and potential solutions. NMR Biomed. 37, e4992. 10.1002/nbm.4992

Patel, J., Shim, I., Agrawal, D.K., 2025. Interventions for Neural Plasticity in Stroke Recovery. Arch. Intern. Med. Res. 8, 246–258. 10.26502/aimr.0217

Pereira, M., Sobolewski, A., Millán, J. del R., 2017. Action Monitoring Cortical Activity Coupled to Submovements. eNeuro 4, ENEURO.0241-17.2017. 10.1523/ENEURO.0241-17.2017

Regularization and Variable Selection via the Elastic Net on JSTOR [WWW Document], n.d. URL https://www-jstor-org.proxy.library.upenn.edu/stable/3647580 (accessed 7.1.26).

Rehab CARES In USA: Clinical Trial | Clinical Research Trial Listing [WWW Document], 2026. URL https://www.centerwatch.com/clinical-trials/listings/NCT05542121/rehab-cares-in-usa-clinical-trial (accessed 7.1.26).

Shahid, J., Kashif, A., Shahid, M.K., 2023. A Comprehensive Review of Physical Therapy Interventions for Stroke Rehabilitation: Impairment-Based Approaches and Functional Goals. Brain Sci. 13, 717. 10.3390/brainsci13050717

Shelton FN, Reding MJ, 2001. Effect of lesion location on upper limb motor recovery after stroke. Stroke 32, 107–112. 10.1161/01.str.32.1.107

Shulman, G.L., Pope, D.L.W., Astafiev, S.V., McAvoy, M.P., Snyder, A.Z., Corbetta, M., 2010. Right Hemisphere Dominance during Spatial Selective Attention and Target Detection Occurs Outside the Dorsal Frontoparietal Network. J. Neurosci. 30, 3640–3651. 10.1523/JNEUROSCI.4085-09.2010

Stinear, C.M., Byblow, W.D., Ackerley, S.J., Smith, M., Borges, V.M., Barber, P.A., 2017. PREP2: A biomarker-based algorithm for predicting upper limb function after stroke. Ann. Clin. Transl. Neurol. 4, 811–820. 10.1002/acn3.488

Vossel, S., Geng, J.J., Fink, G.R., 2014. Dorsal and Ventral Attention Systems. The Neuroscientist 20, 150–159. 10.1177/1073858413494269

Waters, E.L., Azar, R., Opong-Duah, J., Gaardsmoe, S., Johnson, A., Mendonca, R.J., Cacchione, P.Z., Johnson, M.J., 2025. Towards an Optimal Model of Post-Stroke Sensorimotor Control. IEEE Int. Conf. Rehabil. Robot. Proc. 2025, 1313–1319. 10.1109/ICORR66766.2025.11063066

Waters, E.L., Johnson, M.J., 2025. Can an Optimal Control Model Capture Tracking Kinematics? Insights from Healthy and Post-Stroke Robot Interactions. Annu. Int. Conf. IEEE Eng. Med. Biol. Soc. IEEE Eng. Med. Biol. Soc. Annu. Int. Conf. 2025, 1–7. 10.1109/EMBC58623.2025.11252623

Xu, S., Yan, Z., Pan, Y., Yang, Q., Liu, Z., Gao, J., Yang, Y., Wu, Y., Zhang, Yanan, Wang, J., Zhuang, R., Li, C., Zhang, Yongli, Jia, J., 2021. Associations between Upper Extremity Motor Function and Aphasia after Stroke: A Multicenter Cross-Sectional Study. Behav. Neurol. 2021, 9417173. 10.1155/2021/9417173

Yushkevich PA, Gerig G, 2017. ITK-SNAP: An Intractive Medical Image Segmentation Tool to Meet the Need for Expert-Guided Segmentation of Complex Medical Images. IEEE Pulse 8, 54–57. 10.1109/MPUL.2017.2701493

