## Appendix for "Predicting Robot-Based Tracking Performance After Stroke Using Lesion Features and Clinical Assessments: A Pilot Study"

**Appendix A: Image Processing**

For processing CT scans, we followed the following steps:

1.	Convert the selected scan from a DICOM file to a Neuroimaging Informatics Technology Initiative (NIfTI) file using an open-source tool acquired from Chris Rorden

2.	Visually inspect the scans for irregularities

3.	Use the ITK-SNAP manual polygon tool to outline the lesion borders in the axial brain view, comparing different brain views to ensure correct border selection

4.	Save the lesion mask

5.	Wait a week prior to revisiting the sample

6.	Open the scan and mask together and inspect the scan for irregularities again

7.	Once satisfied with the initial mask, open the radiological report that accompanies the scan

8.	Adjust the lesion mask, accordingly, only including areas that look irregular

9.	Save the lesion mask as version 2

10.	A physician was consulted in cases where lesions were difficult to acquire

After generating the lesion masks, we normalized the masks to a general space using a CT-specific template developed [28]. Through this template, CT scans and their corresponding lesion masks were normalized to a standard space that then maps onto the Montreal Neurological Institute (MNI) 152 space. An isotropic voxel size of 1 mm was selected to ensure easy conversion to a milliliter volume. Furthermore, a bounding box with the following dimensions was implemented for scan normalizations:

{90, -126, -72 |

90, 90, 108}

The resulting scans had (181 X 217 X 181) dimensions.

*Atlas Selection*

Although many atlases are available for use, our previous work has shown that the AAL3 may not have the best coverage in subcortical areas [28]. Previously published results analyses showed that when using the Anatomical Atlas 3 (AAL3), the gold standard available atlases, there were indeed subjects with stroke subcortical damage; however, atlas coverage of subcortical areas did not pick up the lesion [28]. To address the need for a whole-brain atlas that maps to the SPM12 space, we utilized the Hammersmith 2003 atlas. The atlas was overlaid on the lesion mask using ITK-SNAP. The volume per region of interest was calculated using ITK-SNAP’s volume and statistics tool, by multiplying the intensity of voxels within a region by the total region volume.

**Appendix B: Simulation of Human Tracking Behavior Based on Feedback Control**

We present a simplified simulation of human tracking behavior at the end effector. This simulation represents how an individual responds to error during robot-based tracking. The simulation model is based on a simple feedback controller with gains on error in position ([Equation]) and velocity ([Equation]). The feedback control gains are fit using least-squares. To better represent the influence of stroke-related motor and cognitive impairments, we modify the target trajectory by introducing bias and delay. The bias constant ([Equation]) is defined as the average of the median of position across all trials. Given that the task has a median position of zero, a median position that is different from zero indicates a bias to one side of the workspace. In simulation, the bias constant is added to the target trajectory, thus shifting the entire perceived target to align with each participant’s bias. The delay constant ([Equation]) is defined as the average lag corresponding to the maximum cross correlation between the participant’s trajectory and the target trajectory. In simulation, the target trajectory is shifted forward in time by this delay value to represent the participant’s delayed movement response.

**Appendix C: Feature Matrix Distribution of Variables**

Figure C.1 shows an example lesion segmentation. Figure C.2 shows the distribution of all potential predictors in the features matrices. As shown in the figure, clinical assessments had more even distributions while lesion volume metrics were left skewed due to a bias towards zero lesion in any given brain region. Presence of diabetes, aphasia, and hemorrhagic stroke were represented as binary variables and showed near even distribution across participants.

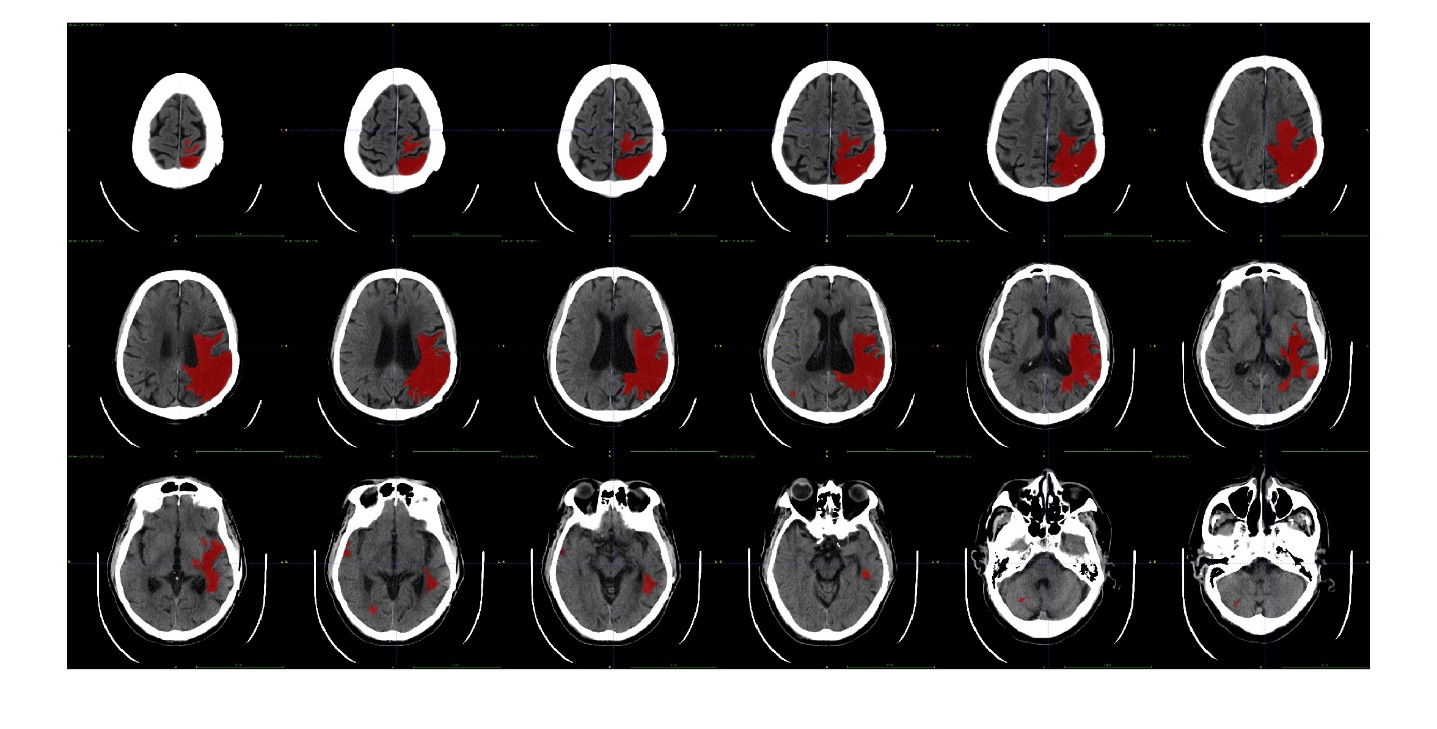
Figure C.1: Subject 05’s lesion development in axial view. The lesioned area is marked in red.

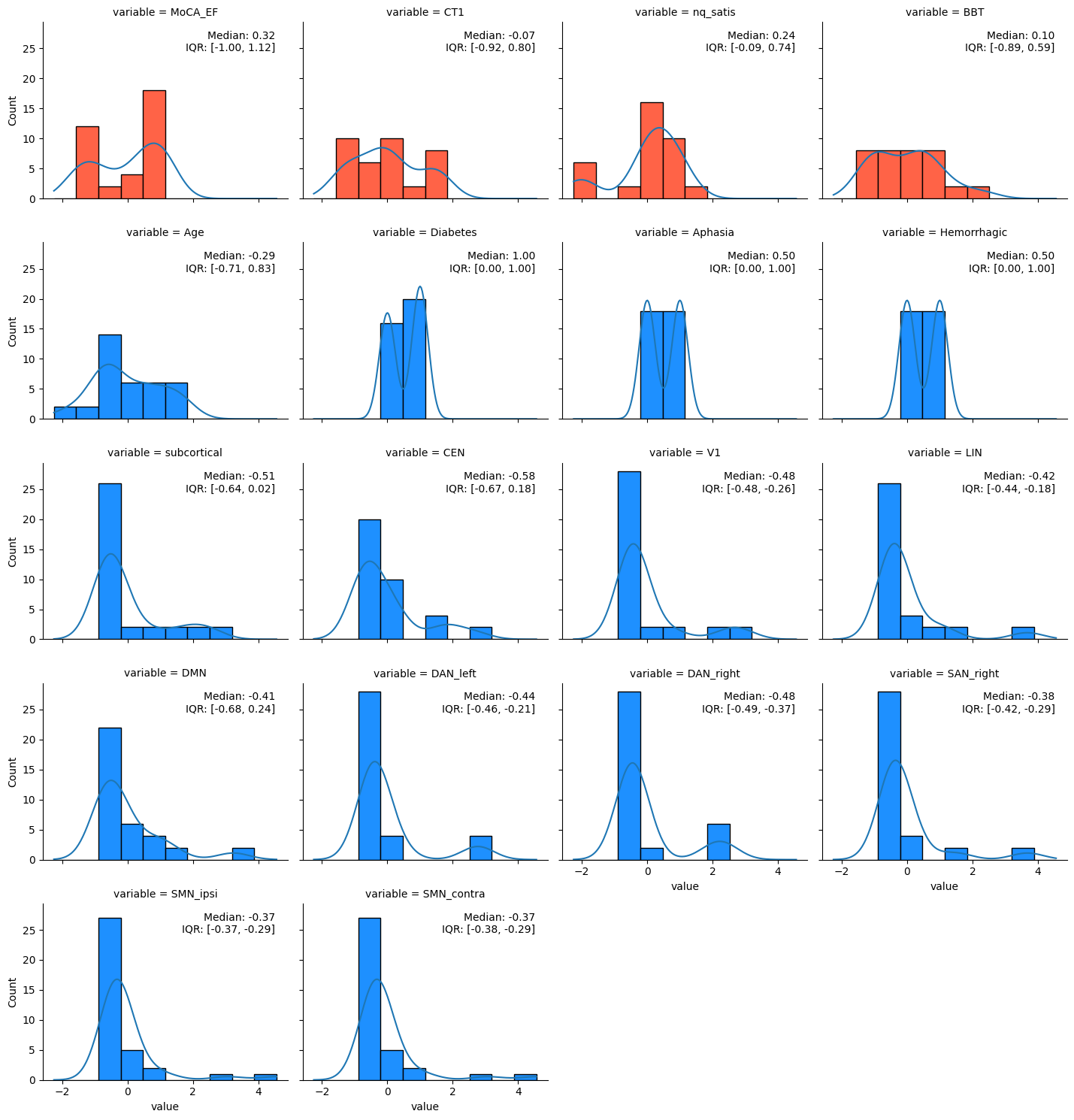
Figure C.2: Distribution of candidate predictors in the feature matrices. Red plots indicate features assigned to the clinical assessment model. Blue plots indicate features assigned to the imaging and clinical outcomes model.

Figure C.3 shows the distribution of kinematic outcomes metrics on the trajectory tracking task for all participants. As shown in the figure, RMSE and ND show even distributions while SPARC has a right bias. For RMSE, a lower value indicates better performance. For ND, a value closer to one indicates that the participant moved as much as the task requires while a value below one indicates that they did not move enough. No participants moved more than the task required (ND > 1). For SPARC, a more negative value indicates less smooth movement.

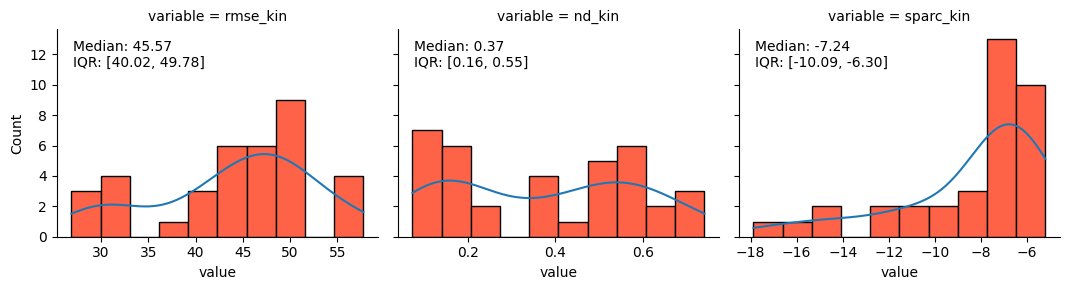
Figure C.3: Distribution of kinematic outcomes on trajectory tracking.

The simulation model accurately captured overall behavior at the end effector (Spearman Rho = 0.45±0.41, Mean Absolute Error = 10.12±3.85 on training data; Spearman Rho = 0.40±0.44, Mean Absolute Error = 10.73±3.65 on testing data). Figure 4 shows example plots for three different participants. Figure 5 summarizes the model parameters for all participants. As shown in the figure, delay and bias display even distributions, while Kv and Kp are left skewed. Because Kp is close to zero for almost all of the participants, it was excluded from the remaining analysis.

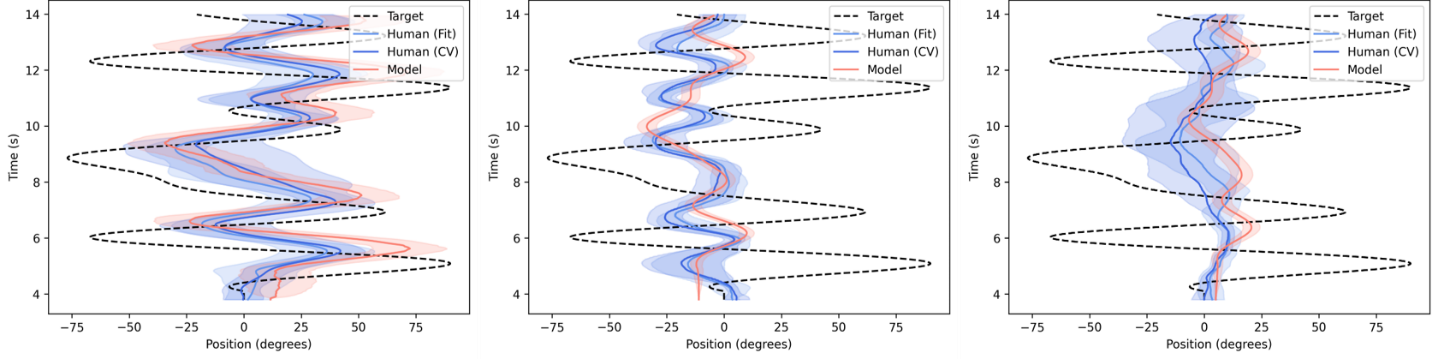
Figure C.4: Example simulation model fits for three participants: 029 left hand (left), 003 left hand (middle), 002 right hand (right).

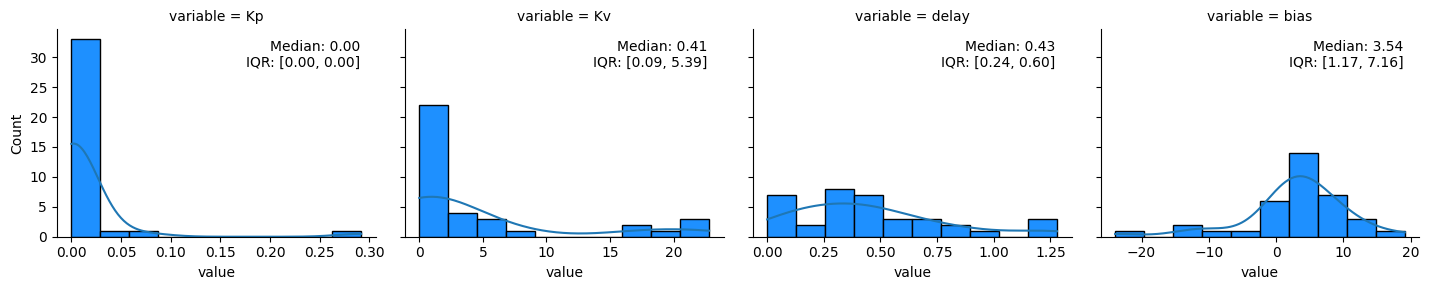
Figure C.5: Distribution of simulation model parameters.

**Appendix D: Stable Features**

Table D.1 shows the stable features for each model, which were defined as features with a selection frequency of 50% or higher.

Table D.1: Stable Features Selected for Each Model

| *Model* | *Metric* | *Feature* | *Selection Frequency (%)* | *Coefficient* |
| --- | --- | --- | --- | --- |
| M3 | RMSE | BBT | 100 | 2.19 ± 0.28 |
|  |  | CT1 | 100 | 1.85 ± 0.22 |
|  |  | Age | 100 | 0.78 ± 0.30 |
|  |  | DAN right | 94 | 0.67 ± 0.25 |
|  | ND | BBT | 100 | 0.05 ± 0.01 |
|  |  | CT1 | 89 | 0.02 ± 0.01 |
|  |  | SAN right | 72 | 0.01 ± 0.01 |
|  | SPARC | BBT | 61 | 0.24 ± 0.24 |
|  |  | MoCA-EF | 50 | 0.18 ± 0.21 |
|  | Delay | DAN right | 100 | 0.09 ± 0.03 |
|  |  | CT1 | 100 | 0.08 ± 0.02 |
|  |  | BBT | 100 | 0.04 ± 0.01 |
|  | Bias | DAN right | 89 | 0.85 ± 0.40 |
|  |  | V1 | 89 | 0.21 ± 0.14 |
|  |  | SMN contra | 83 | 0.15 ± 0.12 |
|  | Kv | CT1 | 100 | 1.44 ± 0.32 |
|  |  | BBT | 100 | 1.00 ± 0.19) |
|  |  | Age | 100 | 0.70 ± 0.23 |
|  |  | Neuro-QoL | 78 | 0.30 ± 0.22 |
| M2 | RMSE | Age | 100 | 1.86 ± 0.36 |
|  |  | DAN right | 94 | 0.63 ± 0.24 |
|  |  | SAN right | 50 | 0.08 ± 0.10 |
|  | SPARC | SMN contra | 61 | 0.13 ± 0.29 |
|  | Delay | DAN right | 94 | 0.08 ± 0.03 |
|  |  | Age | 67 | 0.03 ± 0.02 |
|  | Bias | DAN right | 94 | 1.02 ± 0.35 |
|  |  | V1 | 94 | 0.30 ± 0.14 |
|  |  | SMN contra | 89 | 0.25 ± 0.13 |
|  | Kv | Age | 100 | 1.88 ± 0.39 |
| M1 | RMSE | CT1 | 100 | 2.69 ± 0.22 |
|  |  | BBT | 100 | 2.67 ± 0.26 |
|  | ND | BBT | 100 | 0.06 ± 0.01 |
|  |  | CT1 | 100 | 0.03 ± 0.01 |
|  | SPARC | BBT | 100 | 0.79 ± 0.15 |
|  |  | MoCA-EF | 100 | 0.56 ± 0.10 |
|  | Delay | CT1 | 100 | 0.10 ± 0.01 |
|  |  | BBT | 100 | 0.06 ± 0.01 |
|  | Kv | CT1 | 100 | 2.14 ± 0.31 |
|  |  | BBT | 100 | 1.27 ± 0.18 |
|  |  | Neuro-QoL | 89 | 0.50 ± 0.24 |
